# Modulating delirium through stimulation (MoDeSt): study protocol for a randomized, double-blind, sham-controlled trial assessing the effect of postoperative transcranial electrical stimulation on delirium incidence

**DOI:** 10.1101/2024.07.11.24310269

**Authors:** Sophie Leroy, Viktor Bublitz, Ulrike Grittner, Robert Fleischmann, Falk von Dincklage, Daria Antonenko

## Abstract

- **Background:** Postoperative delirium (POD) is the most common neurological adverse event among elderly patients undergoing surgery. POD is associated with an increased risk for postoperative complications, long-term cognitive decline, an increase in morbidity and mortality as well as extended hospital stays. Delirium prevention and treatment options are currently limited. This study will evaluate the effect of transcranial electrical stimulation (tES) on the incidence of POD.
- **Methods:** We will perform a randomized, double-blind, sham-controlled trial using single-session postoperative application of tES in the recovery room in 225 patients (>65 years) undergoing elective major surgery. Patients will be randomly allocated (ratio 1:1:1) to one of three study groups: (1) alpha-tACS over posterior parietal cortex [2 mA, 20 min], (2) anodal tDCS over left dorsolateral prefrontal cortex [2 mA, 20 min], (3) sham [2 mA, 30 s]. Delirium will be screened twice daily with the 3-minute diagnostic interview Confusion Assessment Method (3D-CAM) in the five days following surgery. The primary outcome is the incidence of POD defined as at least one positive screening during the five first postoperative days compared between tACS and sham groups. Secondary outcomes include delirium severity, duration, phenotype, postoperative pain, postoperative nausea and vomiting, electroencephalographic (EEG) markers, and fluid biomarkers.
- **Discussion**: If effective, tES is a novel, easily applicable, non-invasive method to prevent the occurrence of POD. The comprehensive neurophysiological and biofluid assessments for markers of (neuro-)inflammation and neurodegeneration will shed light on the pathomechanisms behind POD and further elucidate the (after-)effects of tES. The potential implications for the postoperative recovery comprise enhanced patient safety, neurocognitive outcome, perioperative manageability but also reduced healthcare costs.
- **Trial registration**: German Clinical Trial Registry, DRKS00033703, registered on 23 February 2024.

## Introduction

### Background and rationale {6a}

Current demographic scenarios assume that the population over the age of 65 will multiply by three by 2050 and their share of the population will reach 20% (1). Patients in this age group are at particularly high risk of developing postoperative delirium (POD) after elective surgery (2). Clinically, POD manifests as a disturbance of consciousness and attention with an acute onset and fluctuating course. In addition, POD is a risk factor for treatment-associated complications, for long-term cognitive deficits and for the development of care dependency (3–5). Despite these risks, the amount of major surgery in elderly patients is increasing (6). This can be seen impressively in the increase in spinal operations, some of which are being performed up to 28 times more frequently on elderly patients compared to the 1990s (7).

Research findings point to a multifactorial model of delirium pathogenesis that encompasses neural aging, neuroinflammation and endocrine stress responses, resulting in two critical aspects: neuronal neurotransmitter disbalances and network dysfunctions (8,9). These changes are reflected in biofluid and neurophysiological biomarkers (10,11). In particular, perioperative electroencephalogram (EEG) recordings suggest that disconnectivities in the alpha frequency band (8-12 Hz) may play a role in the pathophysiology of POD (12).

The pharmacological treatment of delirium is currently limited to symptomatic measures. The evidence on the benefit of medication is limited and the use of antipsychotics is not a causal treatment of delirium, but only serves to control symptoms (13). Drug therapy with centrally acting drugs is therefore primarily used to dampen overly agitated behaviour and to allow the implementation of necessary therapeutic measures (14). Therefore, various strategies have been developed to prevent POD. Importantly, monitoring intraoperative cerebral activity with a frontal EEG allows individualized adjustment of anaesthetic and analgesic administration to avoid overdosing (15). Optimization of further perioperative factors such as polypharmacy, noise exposure, pain and early postoperative reorientation support can also reduce the incidence of POD (16).

Non-invasive transcranial electrical stimulation (tES), in particular the application of a weak direct (transcranial direct current stimulation, tDCS) or alternating current (transcranial alternating current stimulation, tACS) via surface electrodes attached to the scalp, has shown promising results in the treatment of neurological diseases, including amelioration of age-associated cognitive deficits and chronic pain (17–21). Through the interaction of weak currents with ongoing neural processes, tES has the potential to modulate brain plasticity, neuronal network architecture and cerebral blood flow, thus exerting effects on neural processes that underly pain perception and higher-order cognitive functions (22–24).

During tACS, the oscillating current within a certain physiological frequency range interacts with ongoing cortical activity (25,26). tACS appears to modulate the amplitude, phase, coherence and synchrony of ongoing oscillations and can hereby influence entrainment mechanisms (25–28). This can support interregional synchronization, long-range neuronal coupling and thereby promote successful information transmission, improving cognitive function (29–31). Anodal tDCS over task-relevant brain regions can influence cell membrane potentials and increase cortex excitability by inducing long-term potentiation (LTP)-like processes that outlast the stimulation interval (32). Animal models suggest altered tissue density due to altered neuronal morphology, altered glial cell activity, and reorganization of synaptic connections in terms of neuroplastic phenomena induced by tDCS (33,34). In the only trial to date using tES in patients at risk to develop POD, Tao and colleagues observed a strong effect of postoperative single-session anodal tDCS over the left dorsolateral prefrontal cortex (DLPFC) on the incidence of POD and postoperative pain (35).

### Objectives {7}

The objective of this study is to investigate the effects of tES on the incidence of POD and postoperative pain. As primary objective, the effect of tACS in the alpha frequency range (alpha-tACS) on the incidence of POD after major surgery will be examined in comparison to sham stimulation. In a second study arm, the effect of tDCS compared to sham stimulation on the incidence of POD will be investigated. The effect of tES on postoperative pain will be investigated as a secondary outcome, for both intervention groups in comparison to sham stimulation. To identify potential predictors of the responsiveness to tES and to address the multifactorial genesis of POD and postoperative pain, we plan to assess neuronal network dysfunction with pre-, intra-, and postoperative EEG recordings but also neurotransmitter disbalance and neuroinflammation/- destruction using biofluid blood biomarkers. Furthermore, the influence of intraoperative medication, type and length of surgery and vital parameters on the endpoints will be considered. Potential predictors of response to tES will thus be identified.

### Trial design {8}

This study is a prospective, monocentric, randomized, double-blind, sham-controlled clinical trial. Data collection will occur preoperatively, intraoperatively, and over the first 5 postoperative days. Three trial arms with different types of stimulation intervention will be included: alpha-tACS, tDCS, and sham stimulation with an allocation ratio of 1:1:1.

Patients will undergo a single postoperative tES stimulation (with 2 mA) for 20 minutes in the post-anesthesia care unit (PACU). We will undertake perioperative EEG recordings and blood sampling at three different timepoints. POD and postoperative pain will be screened for one hour in the PACU as well as twice daily for 5 days after surgery.

## Methods: Participants, interventions and outcomes

### Study setting {9}

Single center study of the University Hospital Greifswald (UMG). The study has been approved by the local ethics committee with the registration number BB 179/23 on December 8, 2023.

### Eligibility criteria {10}

#### Inclusion Criteria

- Patients over 65 years of age
- Planned elective general surgery, urological, gynaecological, orthopaedic, or spinal procedures lasting more than 60 minutes (major surgery)
- Capable of providing consent
- Presence of a signed informed consent form

#### Exclusion Criteria

- Known neurological or psychiatric conditions, especially history of epileptic seizures
- Contraindications for tDCS/tACS application (27)
- Long-term medication with central nervous system (CNS) active drugs
- Isolation due to multidrug-resistant germs
- Inability of the patient to read and/or speak German

### Who will take informed consent? {26a}

Screening will take place at the anesthesia outpatient clinic and the inpatient wards of the University Medicine Greifswald during preoperative anesthesia consultations.

After checking the inclusion and exclusion criteria, handing out, and orally explaining the patient information, consent to participate in the study will be requested. Patients will be enrolled after signature of the informed consent form. Subsequently, an identification number will be assigned for pseudonymization.

### Additional consent provisions for collection and use of participant data and biological specimens {26b}

The informed consent form comprises specific sections about the use of biological specimens and patient data. It explains the handling of the biological specimens, stating that the blood samples will be processed using the assigned pseudonyms and stored during the conduction of the trial in the biobank of the University Medicine Greifswald. Subsequently, the intended parameters will be determined, and the blood samples destroyed to ensure data minimization. Informed consent form details the handling of patient data and accords with the General Data Protection Regulation (GDPR).

## Interventions

### Explanation for the choice of comparators {6b}

The choice of comparators in this study, involving alpha-tACS, tDCS and sham stimulation, is grounded in the need to comprehensively evaluate the efficacy of tES protocols in preventing POD. Alpha-tACS and tDCS were selected based on their demonstrated potential in modulating brain activity, which is crucial to address the pathophysiological mechanisms underlying POD. The alpha-tACS intervention was chosen based on the previously described disrupted neural network connectivity in the alpha range associated with POD (36).

tDCS applies a constant electrical current, which can modulate neuronal excitability and has been shown to reduce postoperative pain and the POD incidence when applied postoperatively (35). The sham stimulation serves as the control condition. The sham procedure mimics the application of the active interventions without delivering the electrical current, ensuring that participants, care providers, and outcome assessors are blinded to the intervention.

### Intervention description {11a}

Patients will be enrolled at least one day before surgery. Questionnaires regarding contraindications for tES as well as baseline questionnaires will be handed out (PPI, HADS, STAI, PCS, BRI, PSQ, DN4). The preoperative cognitive status will be assessed with a MoCA score. On the day of the surgery, a 64-channel EEG will be recorded in the preoperative holding area. Preoperative delirium and pain will be assessed with standardized tests (RASS, 3D-CAM and NRS). A preoperative blood sample will be drawn. Before induction of anesthesia a frontal EEG recording will be started that will last until the end of anaesthesia.

A single tES intervention will take place postoperatively in the PACU, after return of consciousness and patient handover to the PACU staff. Emergence of delirium in the PACU (PACU-D) will be assessed, and patients will be asked about postoperative nausea and vomiting (PONV) as well as pain using standardized tests (RASS, 3D-CAM and NRS) every 30 minutes for one hour in the postanaesthetia care unit (PACU). A 64-channel EEG will be recorded, and a second blood sample will be drawn after the tES intervention.

During the first 5 postoperative days, patients will be screened twice daily for POD and pain using standardized tests (RASS, 3D-CAM and NRS). A third blood sample will be drawn on the second postoperative day (Figure 1).

**Figure 1:**
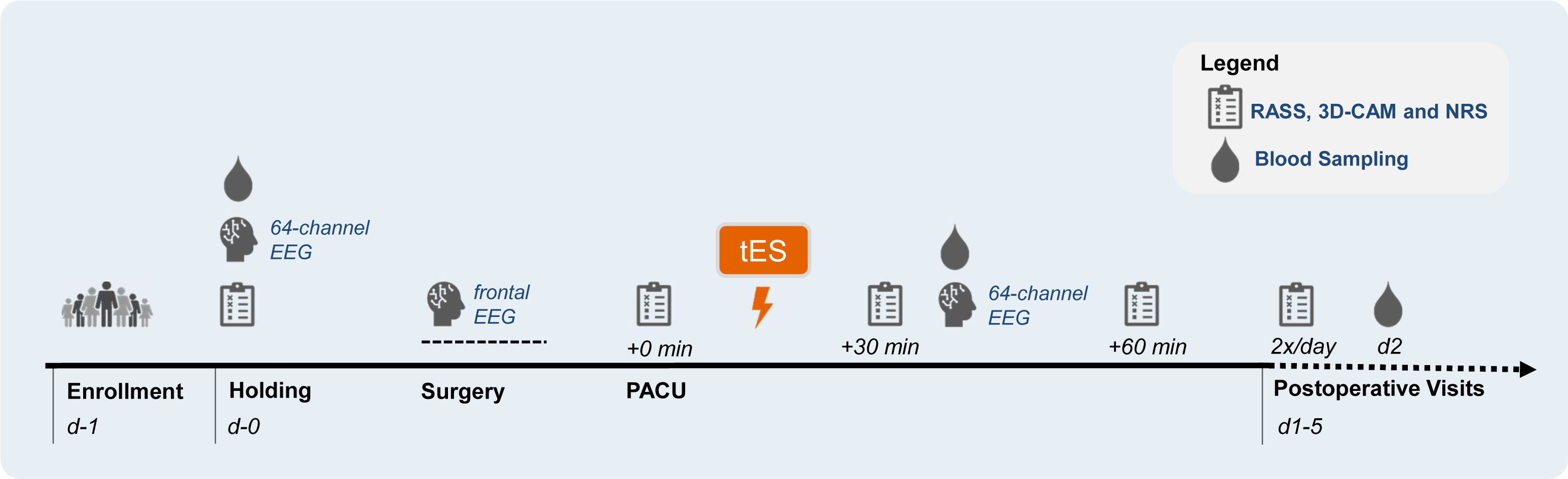
Timeline of the study interventions and visits. PACU: post-anesthesia care unit, EEG: electroencephalography, tES: transcranial electrical stimulation, RASS: Richmond Agitation-Sedation Scale, 3D-CAM: 3-minute diagnostic interview Confusion Assessment Method, NRS: Numerical Rating Scale.

#### Baseline questionnaires

A questionnaire regarding contraindications for the application of tES will be filled out by the physician enrolling the patient. It comprises questions regarding presence of metallic implants, history of neurosurgical intervention, traumatic brain injury or epileptic seizures, known dermatological disorders, ongoing long-term medication, possibility of a pregnancy and previous tES application.

A set of questionnaires regarding pain (NRS, DN4), attitude towards pain (PCS, PSQ) and affective disorders (HASD, STAI) will be explained and handed out to the patient.

The Montreal Cognitive Assessment (MoCA) score evaluates cognitive functioning in areas such as attention, memory, language, and visuospatial skills. It will be conducted and scored by a trained study personnel.

#### EEG

A 5-minute resting-state EEG with eyes closed will be recorded preoperatively in the holding area and post-intervention in the PACU. EEG data will be recorded using ANT Neuro’s dry EEG system, equipped with dry surface electrodes that do not require the use of conductive gel or paste. This system is designed for a rapid setup, allowing for signal acquisition in this clinical setup. Patients will be lying in bed, and the dry EEG cap, fitted with 64 electrodes according to the 10-20 international system, will be placed on their head. The electrode positions will be adjusted to ensure optimal scalp contact. Patients will be asked to remain still and relaxed, with eyes closed during the recording session of 5 minutes. Data acquisition will be conducted at a sampling rate of 1064 Hz. The impendences will be monitored and maintained below an ideal threshold of 5 kΩ if possible or the recommended threshold of 100 kΩ.

Intraoperatively, a frontal EEG will be recorded with a common neuromonitoring tool available at the study site (SedLine® Brain Function Monitor by Masimo or Narcotrend®-Compact M by Narcotrend-Gruppe). Both medical devices are used to monitor the brain function of patients under anesthesia or sedation in critical care settings, utilizing raw and quantitative EEG parameters to provide a real-time dimensionless index of sedation depth. Before the start of the recording the study staff ensures the proper functioning of the monitor. For the SedLine Monitor, we will make sure that the feed speed is set at 30 mm/s and the amplitude at 5 mV, since the display settings affect the sampling frequency of the recorded data stored (37).

#### Intervention

The tES will be administered using a battery-powered stimulator (Neuroelectrics® Starstim Home Research Kit, Barcelona, Spain) via two electrodes (5 cm in diameter) soaked with 0.9% NaCl solution and mounted in a neoprene cap.

To ensure blinding to the stimulation group, the ECG monitoring in the PACU will be masked before the start of the intervention with a stencil covering the raw ECG traces.

In the tACS group, two electrodes will be placed at the P3 and P4 positions of the 10-20 EEG system, over the respective left and right posterior parietal cortex (PPC) (Figure 2B). A 20-minute oscillating stimulation with 2 mA (peak-to-peak) at an age-adapted frequency of 9.5 Hz will be applied. An age-adapted stimulation frequency was determined for this cohort by analysing a database of 532 clinical resting-state physiological EEG recordings. The method was described in detail elsewhere (38). In the tDCS group, the anode will be placed over the left DLPFC, as determined by the F3 position of the 10-20 EEG system. The cathode will be placed over the right supraorbital area, corresponding to Fp2 of the 10-20 system (Figure 2A). The 20-minute continuous stimulation with 2mA will gradually increase and decrease at the beginning and end within an additional ten second period (ramp time).

**Figure 2:**
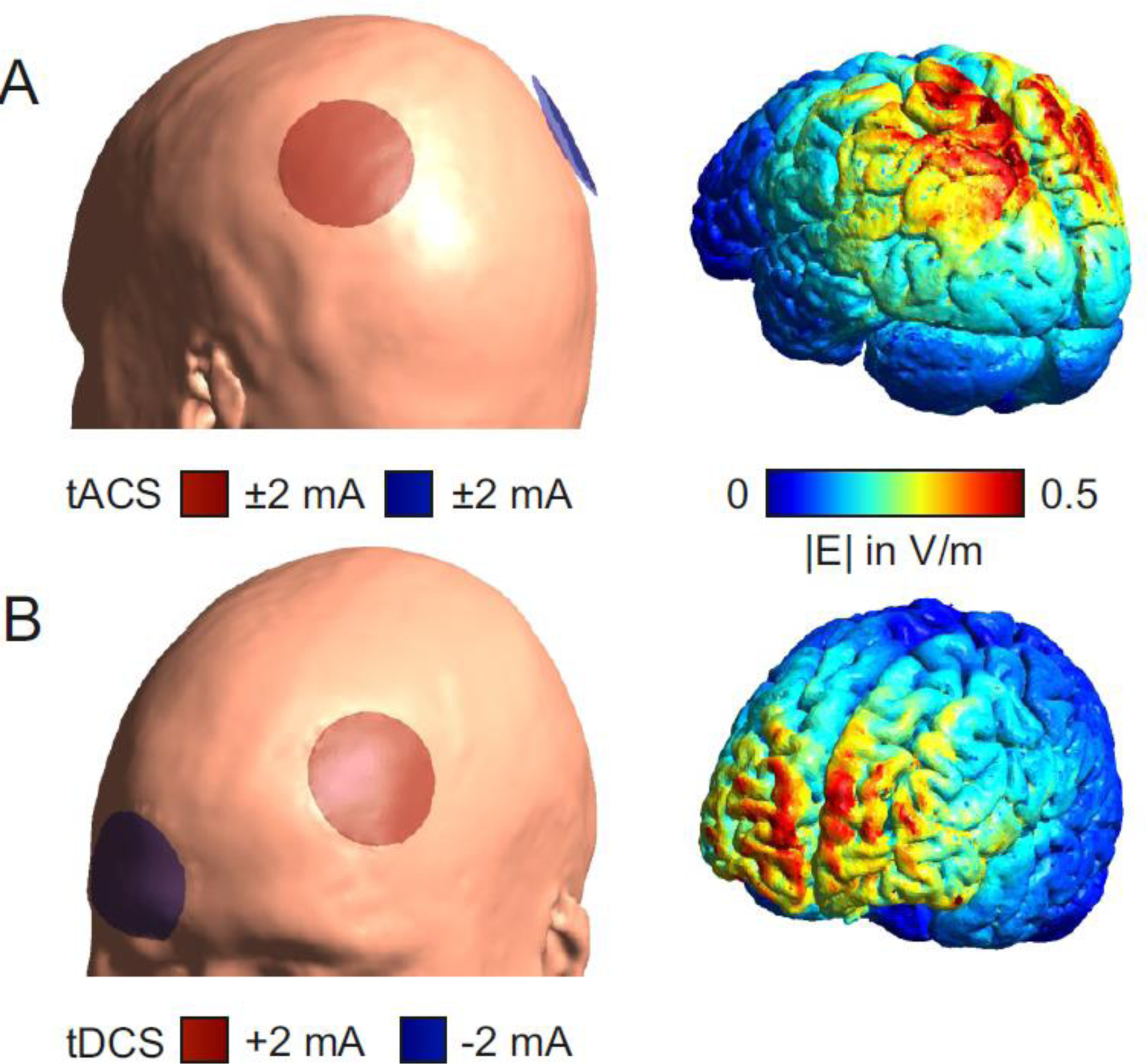
Illustration of electrode placement and electric field distribution. Applied transcranial electrical stimulation on an MNI head/brain for (A) tACS group (stimulation frequency: 9.5 Hz; electrodes centered over P3 and P4; electrode diameter: 5 cm) and (B) tDCS group (electrodes centered over F3, anode, and Fp2, cathode; electrode diameter: 5 cm), simulated using SimNibs (39,40). tACS: transcranial alternating current stimulation, tDCS: transcranial direct current stimulation.

The sham group is divided into two subgroups with n(tDCS)/2 for tDCS-sham and n(tACS)/2 for tACS-sham. In these subgroups, the same electrode arrangements and ramp times are used, but the current is only applied for 30 seconds to induce the typical tingling sensation on the scalp and blind the patients to the stimulation condition.

The perception of side effects associated with the stimulation and group affiliation is surveyed at the end of each stimulation condition using a standardized questionnaire (27).

#### Blood samples

Three blood samples will be taken for the study: one preoperatively, one postoperatively in the PACU, and one on the second postoperative day, with 5 mL of serum collected each time. The blood samples will be sent immediately after collection to the University Medicine Greifswald biobank for aliquoting and storing. The following parameters will then be determined from these samples in the neurology research laboratory of the University Medicine Greifswald: VILIP-1, CCL2 (MCP-1), sTREM-2, BDNF, TGF-β1, VEGF, IL-6, sTREM-1, β-NGF, IL-18, TNF-α, sRAGE, CX3CL1, CRP, NfL, S100b, VCAM-1, ECAM-1. The samples will be destroyed after measurement.

#### Postoperative visits

The RASS, the 3D-CAM and the NRS score will be assessed once preoperatively in the holding area, every 30 minutes for one hour in the PACU (3 times in total) as well as twice a day for five days postoperatively.

The Richmond Agitation-Sedation Scale (RASS) score is a commonly used tool to assess a patient’s level of agitation or sedation, ranging from +4 (combative) to -5 (unarousable), with 0 representing an alert and calm state (41). Patients with a postoperative RASS score in the PACU of -2 or below will not be questioned regarding POD or pain.

The 3D-CAM is a reliable, validated diagnostic assessment tool for delirium, based on a 3-minute interview to identify the presence of POD (42). Based on the principle of the CAM method,” (…) the algorithm is considered positive if following features are present: Feature 1) Acute onset or fluctuating course and Feature 2) Inattention and either Feature 3) Disorganized thinking or Feature 4) Altered level of consciousness”.

If a patient is identified as delirious based on the 3D-CAM, the RASS score will be used to determine the phenotype of the delirium at the timepoint of the visit: hypoactive (RASS -5 to -1) or hyperactive (RASS +1 to +4).

Furthermore, the 3D-CAM-S score will be calculated from the 3D-CAM assessment. The 3D-CAM-S is a validated method derived from the 3D-CAM diagnostic interview for assessing delirium severity, demonstrating excellent agreement with the commonly used delirium severity scale, CAM-S Short Form, and is designed to provide prognostic information, monitor delirium progression, and evaluate treatment response in older patients undergoing major elective non-cardiac surgery (43).

The Numeric Rating Scale (NRS) is a self-report measure of pain intensity on a scale of 0 (no pain) to 10 (worst possible pain), used for assessing a patient’s self-reported pain level.

### Criteria for discontinuing or modifying allocated interventions {11b}

Patients may prematurely drop out of the study under the following conditions:

- The personal wish of the patient
- The occurrence or discovery of events or criteria after enrolment that contradict

intervention.

A premature end of the study or termination of the entire study may be considered under the following circumstances:

- New findings during the clinical trial that could endanger the safety of the participants (positive benefit-risk assessment no longer given).

### Strategies to improve adherence to interventions {11c}

n./a.

### Relevant concomitant care permitted or prohibited during the trial {11d}

Apart from the intervention and the close monitoring for POD during the postoperative visits, the participation in the study will not further affect the care of patients. Concerning the perioperative care, the anesthetic management protocols will not be modified by the trial. Induction and maintenance of anesthesia will be conducted according to the standard operating procedure of the clinic. The treating anaesthesiologist will have access to the routinely used neuromonitoring devices and the indices of depth of anesthesia. If patients develop a POD, the treatment of productive delirious symptoms with antipsychotic drugs will be administered as prescribed by the treating ward physician.

At the University Hospital Greifswald, a delirium prevention unit has been implemented in the routine care of hospitalized patients of four departments (general surgery, orthopedy, internal medicine and neurology). All patients over 65 years of age are screened daily by the unit, and non-pharmacological delirium prevention interventions are undertaken. Patients who are enrolled in the trial and admitted to one of these departments will not be visited or accompanied by the delirium prevention unit.

### Provisions for post-trial care {30}

n/a

No provisions for ancillary and post-trial care are expected to be relevant to this study.

### Outcomes {12}

#### Primary Outcome Measure

Incidence of POD as defined by having at least one positive 3D-CAM within the 10 visits in the 5 postoperative in-hospital days.

#### Secondary Outcome Measures

- Duration of POD (days between first positive 3D-CAM score and last positive 3D-CAM
score)
- Maximal POD severity (3D-CAM-S)
- Median and interquartile range of POD severity (3D-CAM-S)
- Delirium phenotype (RASS)
- Adverse events
- Incidence of emergency delirium in the PACU (PACU-D) as defined by having at least one positive 3D-CAM score in the PACU
- Postoperative pain in the PACU (NRS)
- Postoperative pain in the five first postoperative days (NRS)
- Presence of postoperative nausea and vomiting (PONV)
- EEG biomarkers from postoperative 64-channel EEG recordings including but not limited to: total power, power within frequency bands, measures of periodic activity (power, center frequency, bandwidth), measures of aperiodic activity (offset, exponent), phase-lag-index, entropy measures, burst-suppression-ratio.
- Postoperative blood biomarkers

#### Additional covariates

- Preoperative pain, attitude towards pain and affective disorders (PPI, HADS, STAI, PCS, BRI, PSQ, DN4).
- Preoperative cognitive score (MoCA)
- EEG biomarkers from pre- and intraoperative EEG recordings
- Preoperative blood biomarkers
- Intraoperative data related to anesthesia: anesthesia management, vital parameters, complications.
- Surgical data: type of surgery, duration of surgery, complications.
- Clinical treatment data: demographics, medical history, sensory impairments (vision aids, hearing aids), medication history, preoperative substance abuse, intensive care unit stay, perioperative transfusions, complications, number and type of lines (e.g., intravenous lines, central venous catheters, arterial blood pressure monitoring, nasogastric or percutaneous feeding tube, suprapubic or urinary catheter, possibly other drains), discharge/transfer destination, length of stay.

### Participant timeline {13}

Patients will receive one session of tES and will be visited postoperatively for 5 days (Table 1).

**Table 1:** Schedule of intervention and visits.

| Timepoint | Measurement | Visits |  |  |  |  |
| --- | --- | --- | --- | --- | --- | --- |
|  |  | V0<br>D-1 | V1<br>H-1 | V2<br>H+0 | V3<br>H+1 | V4-V13<br>D+1 to D+5<br>twice daily |
| Time relative to surgery (D: day, H: hour) |  |  |  |  |  |  |
| Enrollment |  |  |  |  |  |  |
|  | Eligibility screening | x |  |  |  |  |
|  | Informed consent | x |  |  |  |  |
| Preoperative assessments |  |  |  |  |  |  |
|  | Demographic data | x |  |  |  |  |
|  | Medical History | x |  |  |  |  |
|  | Baseline questionnaires | x |  |  |  |  |
|  | Cognitive status (MoCA) | x |  |  |  |  |
| Intervention |  |  |  |  |  |  |
|  | Alpha-tACS / tDCS / sham |  |  |  | x |  |
| Postoperative assessments |  |  |  |  |  |  |
|  | Arousal (RASS) |  | x |  | x | x |
|  | POD (3D-CAM) |  | x |  | x | x |
|  | Pain (NRS) |  | x |  | x | x |
| Physical measures |  |  |  |  |  |  |
|  | 64-channel resting-state EEG recording (5 minutes) |  | x |  | x |  |
|  | Intraoperative frontal EEG recording |  |  | x |  |  |
|  | Blood draw |  | x |  | x | x |
|  |  |  |  |  |  | only D+2 morning |
| Abbreviations: MoCA: Montreal Cognitive Assessment. tACS: transcranial alternating current stimulation. tDCS: transcranial direct current stimulation. RASS: Richmond Agitation-Sedation Scale. 3D-CAM : 3-minute diagnostic interview Confusion Assessment Method. NRS: Numerical Rating Scale. |  |  |  |  |  |  |

### Sample size {14}

For the effect of tDCS on the frequency of postoperative delirium, the trial by Tao and colleagues observed a reduction from 19.7% to 4.9% (35) (Odds ratio: 4.761). A previous study on the incidence of POD among general surgical patients at University Medicine Greifswald showed a prevalence of 22.2% (42). We assume an effect similar to those of Tao et al. To demonstrate such an effect between the tACS-group and the sham-stimulation group using a Chi-Square-Test with a power of 80% and a one-sided significance level alpha of 0.05, a group size of N=60 is required. With 3 groups, and an expected dropout rate of about 20%, in total N=225 participants should be included (Figure 3).

**Figure 3:**
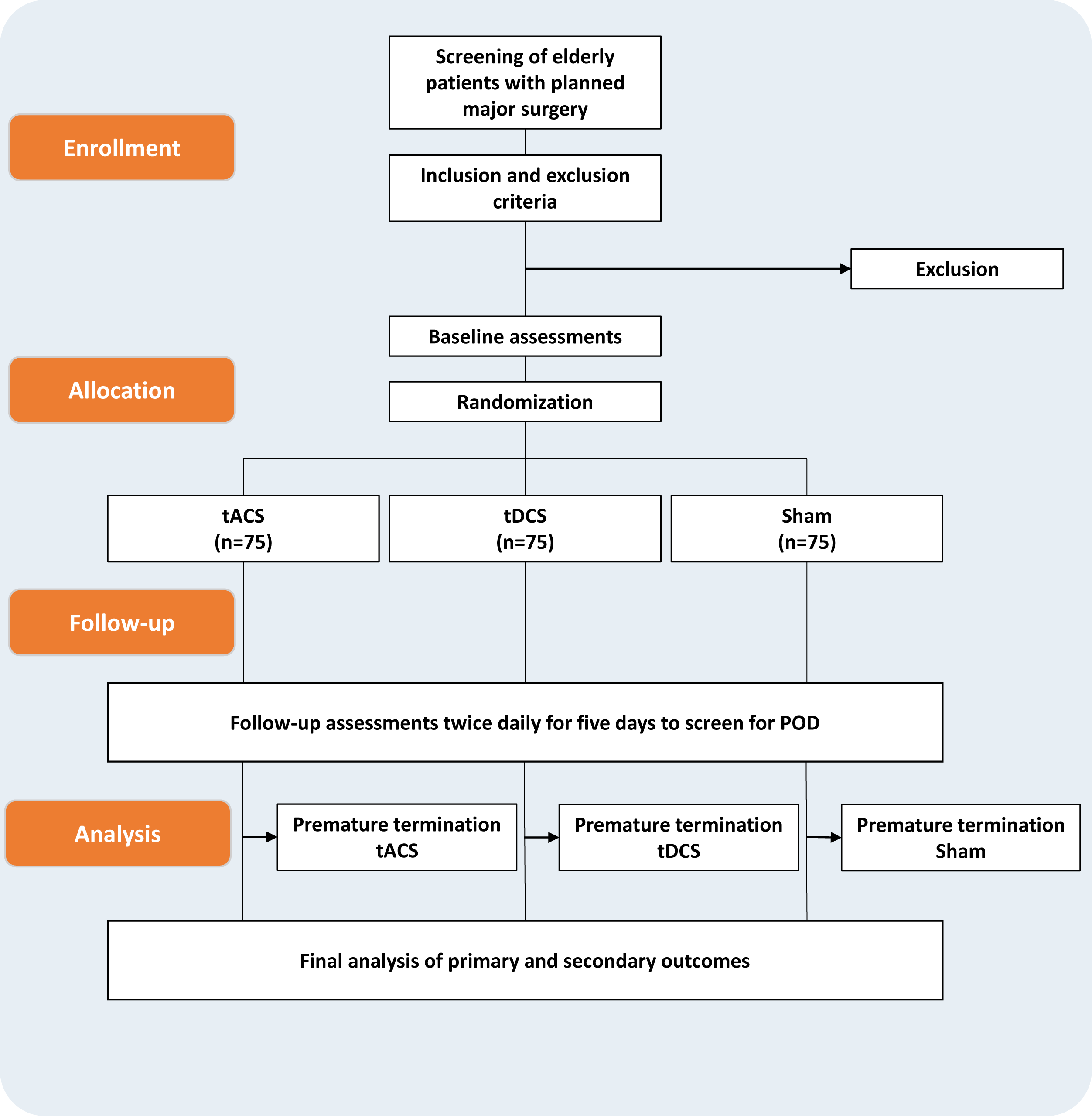
Flow-chart of the study processes. tACS: transcranial alternating current stimulation, tDCS: transcranial direct current stimulation.

### Recruitment {15}

To achieve adequate participant enrolment, patients eligible for the trial will be screened daily by VB and SL and all patients meeting all the inclusion criteria and no exclusion criteria will be asked to participate in the trial. Due to the large number of operations conducted daily in the departments of interest at University Hospital Greifswald we do not expect problems with the recruitment of patients.

## Assignment of interventions: allocation

### Sequence generation {16a}

Assignment to the alpha-tACS, tDCS, and sham groups is done through stratified block randomization. Before the start of the study, a randomization sequence was created by a scientist who is not involved in the project. Using the R software (http://www.R-project.org), randomization blocks of different sizes were created. Participants will be then assigned to the tDCS, tACS, or sham group based on the generated randomization sequences within each block and stratum. The allocation to the experimental groups was done in a 1:1:1 ratio, with age (65-72 years and ≥73 years) and sex (men and women) serving as strata. Since the sham-group was divided in n/2 tACS-sham and n/2 tDCS-sham, the intervention groups were also divided by two and a total of 6 groups were formed: 2x tACS, 2x tDCS, 1x sham-tACS, 1x-sham-tDCS. This repartition ensures that all groups are the same size, and that the frequency of an allocation will not allow tracing back to the intervention group.

### Concealment mechanism {16b}

A randomization sequence was created without allocation to a pseudonym or any specification for each stratum. Patients will be allocated to their group within the stratum following their order of enrolment.

### Implementation {16c}

The enrolment of patients and assignment to the groups is performed by two investigators (VB and SL) who have no access to the allocation sequence generated by a scientist who is not involved in the project.

## Assignment of interventions: Blinding

### Who will be blinded {17a}

Everyone directly involved in this trial will be blinded to the interventions: patients, study staff and care providers outside the study group. The analysis of the primary outcome will be undertaken before unblinding of the allocation sequence.

### Procedure for unblinding if needed {17b}

An Emergency Code Break was provided to the principal investigators, who can open it in emergency situations.

## Data collection and management

### Plans for assessment and collection of outcomes {18a}

#### Questionnaires

We will ask patients to report their pain level during rest and movement and the resulting disturbance on a Numerical Rating Scale from 0-10 (NRS) on the present day. We will furthermore ask patients to report the expected pain level in the PACU and six months after surgery during rest and movement. The Hospital Anxiety and Depression Scale (HADS) is a tool to detect states of depression and its severity (44). The German adaptation has been validated (45). The State-Trait Anxiety Inventory (STAI) score measures two types of anxiety: state anxiety, reflecting temporary feelings of anxiety, and trait anxiety, indicating a more general and long-term level of anxiety. The Pain Catastrophizing Scale (PCS) score measures the degree of catastrophizing thoughts and feelings individuals have about their pain. The Pain Sensitivity Questionnaire (PSQ) score quantifies an individual’s sensitivity to pain based on their responses to various hypothetical pain scenarios. The DN4 (Douleur Neuropathique en 4 Questions) questionnaire is a diagnostic tool used to identify neuropathic pain, consisting of 10 items that assess clinical symptoms and sensory signs.

#### Laboratory Tests

Our study protocol mandates duplicate measurements for ELISA tests to bolster data quality. This practice aims to enhance precision, allow for the identification and correction of any technical errors, and ensure the reliability of our outcomes. We will use the LEGENDplex™ Human Neuroinflammation Panel 1 (13-plex) as well as single parameter ELISA for further specific biomarkers of interest.

### Plans to promote participant retention and complete follow-up {18b}

To improve the compliance, patients will be well-informed about the intervention and the postoperative visits. Patients are more likely to adhere to the trial protocol and complete all trial visits if they are thoroughly informed about the possible benefit and the very low risk associated with the intervention. Due to the short duration of the trial with a single tES application and the in-hospital context of the visits we do not expect major issues with losses to follow-up.

If patients are discharged before the end of the postoperative observation period of five days they will not be considered as loss to follow-up, as a stable neurocognitive outcome is considered as indispensable for patient discharge.

### Data management {19}

This trial was approved by the data protection officer of the University Hospital Greifswald. Clinical data will be collected in a pseudonymized form in a paper case report form (pCRF) for each patient. The digitalization into an electronic case report form (eCRF) will occur with a dual control principle to ensure minimization of transcription errors. Intraoperative EEG data will be collected anonymously, the assignment of the raw EEG traces to the patients will occur based on the date and time of the recording. The 64-channels EEG will be recorded with the ANT Neuro software using pseudonyms on a computer reserved solely for study purposes. Data will be kept for 10 years before being deleted or destroyed.

### Confidentiality {27}

Digital source data are stored on a study-specific, clinic-internal network drive with the corresponding pseudonym. Study leaders ensure through network permissions that only authorized study personnel have access. The allocation list of the pseudonyms will be kept in a locked cabinet.

The responsibility of transferring clinical study data to project partners rests with the aforementioned project leaders. If required, the collected study data may be shared with project partners within the scope of the GDPR, who can demonstrate a suitable data protection concept, for answering study-specific questions, in a pseudonymized form. The data will be published exclusively in an anonymized and aggregated form.

### Plans for collection, laboratory evaluation and storage of biological specimens for genetic or molecular analysis in this trial/future use {33}

The serum samples will be stored in the biobank of the University Medicine Greifswald until the end of the trial when all samples will be analysed collectively. The aforementioned parameters will be determined. If recent scientific findings point towards new biomarkers of interest the investigators leave themselves the option open to add those to the intended parameters.

## Statistical methods

### Statistical methods for primary and secondary outcomes {20a}

The statistical analysis will be based on the intention-to-treat principle, including all randomized patients in the groups to which they were assigned, regardless of the treatment received. To address missing data, we will employ a two-pronged approach. For missing values assumed to be Missing Completely at Random (MCAR) or Missing at Random (MAR), we will utilize Multiple Imputation by Chained Equations (MICE). This method iteratively imputes missing values based on the relationships between variables, resulting in 30 complete datasets for subsequent analyses. In cases where the reason for missingness is known (informed missingness), targeted single imputation techniques will be employed. The specific imputation strategy (worst-case or best-case scenario) will be chosen based on the nature of the missing data. This approach mitigates the effects of dropouts or deviations from the protocol.

#### Primary Outcome Analysis

- Binary logistic regression will be employed to adjust for baseline delirium status and to test differences between treatment (tACS) and control (sham) in incidence of POD. Dependent variable will be incidence of POD (yes/no), independent variables are group (tACS/sham) and baseline delirium status as measured by the 3D-CAM score. Odds ratio and corresponding 95%-CI will be reported.

#### Secondary Outcome Analysis

- Binary logistic regression will be employed to adjust for baseline delirium status and to test differences between treatment (tDCS) and control (sham) in incidence of POD. Dependent variable will be incidence of POD (yes/no), independent variables are group (tDCS/sham) and baseline delirium status as measured by the 3D-CAM score. Odds ratio and corresponding 95%-CI will be reported.
- Continuous variables (e.g., duration of delirium, severity scores, EEG biomarkers, fluid biomarkers) will be analysed using analysis of covariance (ANCOVA) including the particular baseline value.
- Categorical variables (e.g., delirium phenotype) will be analysed using binary logistic regression, adjusting for baseline 3D-CAM.
- For EEG and biofluid biomarker outcomes, linear mixed-models (LMM) will be implemented to account for within-subject correlations across time points.

### Interim analyses {21b}

n./a.

We do not plan any interim analyses.

### Methods for additional analyses (e.g. subgroup analyses) {20b}

Subgroup analyses will be conducted to explore the differential effects of alpha-tACS and tDCS in specific patient subsets, such as by age, sex, type of surgery, or baseline cognitive function.

Subgroup analyses will be conducted by using interaction terms of subgroup by treatment group in the particular regression models.

### Methods in analysis to handle protocol non-adherence and any statistical methods to handle missing data {20c}

The approach to handling missing data will be outlined, with imputation methods, depending on the nature and extent of the missingness.

### Plans to give access to the full protocol, participant level-data and statistical code

**{31c}**

The contact of the corresponding author will be communicated allowing researchers to get in touch with the investigators. Data can be made available upon written request in conformance with the GDPR.

## Oversight and monitoring

### Composition of the coordinating center and trial steering committee {5d}

The trial is coordinated by a team composed of senior and junior neurologists, anaesthesiologists and psychologists. The principal investigators will oversee and train one junior physician from the department of anaesthesiology and one junior physician from the department of neurology who will be running the trial day-by-day and either assist or oversee the intervention. A monthly meeting will take place to discuss the trial status as well as issues that occur.

### Composition of the data monitoring committee, its role and reporting structure {21a}

There will be no independent data monitoring committee due to the lack of external sponsors.

### Adverse event reporting and harms {22}

TES allows for painless modulation of cortical activity and excitability through the intact skull. Within the application parameters described above, tES is proven to be a well-tolerated technique with few, mild side effects. At the standard parameters of tES (intensity 2 mA), a meta-analysis described the primary undesirable side effects as itching (39%), tingling (22%), headaches (15%), burning (9%), and discomfort (10%) (46). All side effects were mild and short-lived, with most symptoms showing no difference between active stimulation and sham stimulation (47). Rarely, and only after hours of continuous use, mild headaches and skin irritation at the electrode sites occur, which completely subside after the end of use.

However, we will closely monitor adverse events during and after the intervention. Concretely, study staff will be continuously present during the intervention. A physician will always assist to the intervention in the PACU or be on call in-hospital. Due to the setting in the PACU, physicians are permanently present in the room in case of adverse effects occurring. After the intervention patients will be asked about itching, pain, burning, feeling of warmth/heat, metallic iron taste, fatigue/reduced attention, or other symptoms. Any adverse event will be documented and immediately reported to the principal investigator.

### Frequency and plans for auditing trial conduct {23}

n./a.

We do not plan to audit the trial.

### Plans for communicating important protocol amendments to relevant parties (e.g. trial participants, ethical committees) {25}

All protocol amendments will be communicated to the local ethics committee of the University Hospital Greifswald. The registry entry will be updated, as well as the published trial protocol.

### Dissemination plans {31a}

The results of the study will be published in peer-review journals in the interest of disseminating scientific knowledge, irrespective of the result.

## Discussion

The aim of this trial is to assess the effectiveness of a single postoperative transcranial electrical stimulation (tES) session in the PACU to prevent postoperative delirium (POD) in elderly patients undergoing elective major surgery. Interventions will compare alpha-tACS (9.5 Hz, 2mA peak-to-peak, 20 minutes) over the PPC and anodal tDCS (2 mA, 20 minutes) over the left DLPFC with sham.

With the MoDeSt trial, we address the growing need for safe treatment protocols for elderly patients when surgery in general anesthesia is necessary. The prevention of POD and postoperative pain by tES poses a potential way to alleviate a relevant burden a surgical intervention can impose on elderly patients. This may prove an essential approach to avoid the emergence of the long-term neurocognitive decline associated with POD, leading to severe consequences for individuals as well as profound socioeconomic impacts.

Research in the neuroanaesthetic field has extensively shown the critical association between POD and pre-(48), intra- (11,49) and postoperative (50–52) EEG power in the alpha band. Low intraoperative alpha power seems to reflect the EEG phenotype of a “vulnerable brain” (52,53). Interventions modulating this neurophysiological characteristic were lacking. A promising approach is to maximize alpha power with intraoperative pharmacological manipulation (54). In the alpha-tACS arm of this trial we will investigate if the stimulation with a weak alternating current in the alpha frequency range during emergence of anesthesia modulates the neural network communication and thereby influences the postoperative neurocognitive outcome.

Our comprehensive outcome measures, including the incidence, phenotype and severity of delirium, alongside complex neurophysiological and biofluid biomarker evaluations, will offer an in-depth insight into the impact of tES, but will also shed light upon the neurophysiological background of POD. Application of tES has been associated with reduced levels of inflammatory biomarkers (IL-6, IL-10, TNF-alpha, ß-endorphin, and BNDF) commonly linked with POD (55,56), though the impact of postoperative tES on perioperative biomarker dynamics has yet to be examined. Developing predictive models based on perioperative clinical data and biomarkers will potentially identify patients who could most benefit from tES interventions. If effective, tES could become a simple, easy-to-use and cost-efficient method to enhance neurocognitive recovery after general anesthesia.

Overall, the trial’s outcomes will explore the therapeutic potential of postoperative tES in elderly patients undergoing general anesthesia for major surgery and contribute to our understanding of the pathophysiological correlates of delirium.

## Trial status

This is the first version of the protocol, dated 02 April 2024. Recruitment began in February 2024 and is expected to be completed within one year. Up to date (April 2024) 28 patients completed the trial.

## Abbreviations

POD: postoperative delirium
tES: transcranial electrical stimulation
tDCS: transcranial direct-current stimulation
tACS: transcranial alternating-current stimulation
EEG: electroencephalography
GDPR: general data protection regulation
PONV: postoperative nausea and vomiting
DLPFC: dorsolateral prefrontal cortex
PPC: posterior parietal cortex.

## Declarations

## Data Availability

All data produced in the present study are available upon reasonable request to the authors.

## Acknowledgements

Not applicable

## Authors’ contributions {31b}

DA conceived the experimental study hypotheses. SL, VB, UG, RF and FD developed the detailed clinical trial protocol. VB and SL are implementing the trial, performing recruitment and supervising the conduct. SL and DA wrote the first draft of the manuscript. All authors read and approved the final version of the manuscript.

## Availability of data and materials {29}

The datasets will be made available from the corresponding author upon written reasonable request.

## Ethics approval and consent to participate {24}

The study has been approved by the local ethics committee with the registration number BB 179/23 on December 8, 2023.

## Consent for publication {32}

Not applicable

## Authors’ information (optional)

n./a.

